# Management of patients with occupational burnout in Switzerland: some insights on the heterogeneity in current practices

**DOI:** 10.1101/2023.11.10.23298381

**Authors:** Irina Guseva Canu, Roger Getzmann, Yara Shoman, Fulvia Rota, Stéphane Saillant, Roland von Känel, Christine Cohidon, Catherine Lazor-Blanchet, Lysiane Rochat, Rafaël Weissbrodt, Nadia Droz, Anny Wahlen

**Affiliations:** Department of Environmental and Occupational Health, Center for Primary Care and Public Health (Unisanté), University of Lausanne, Lausanne, Switzerland; Swiss Society for Psychiatry and Psychotherapy, Bern, Switzerland; Centre Neuchâtelois de Psychiatrie (CNP), Neuchatel, Switzerland; University of Lausanne, Lausanne, Switzerland; Department of Consultation-Liaison Psychiatry and Psychosomatic Medicine, University Hospital Zurich, University of Zurich, Switzerland; Department of Family Medicine, Center for Primary Care and Public Health (Unisanté), University of Lausanne, Lausanne, Switzerland; Occupational Health Service, Lausanne University Hospital, Switzerland; School of health sciences, HES-SO Valais-Wallis, Sion, Switzerland; PSY4WORK.ch, the Swiss Association of work & organization psychologists, Lausanne, Switzerland

**Author notes:** Corresponding author Prof. Guseva Canu Irina Department of Environmental and Occupational Health, Unisanté, University of Lausanne 2, Route de la corniche, 1066 Epalinges ORCID: 0000-0001-7059-8421. Equally contributing authors (Nadia Droz, Anny Wahlen).

**Keywords:** occupational burnout, therapy, severity stage, relapse, return to work

## Abstract

We aimed to describe the characteristics and current practices of Swiss health professionals who manage patients with occupational burnout (POB), namely the general practitioners (GP), psychiatrist-psychotherapists (PP), occupational physicians (OP) and psychologists. Among 3216 respondents, 2951 reported to consult POB, and 1130 (713 physicians and 410 psychologists) to treat them. The study showed that POB management constitutes 5 to 25% of health care professionals’ consultations, with an inequal distribution of POBs across professionals’ specialties and specializations, but also across geographic regions. The profile of POB consulted also differs across professionals. Work psychologists see more often POB at early burnout stage, GPs have most patients with moderate burnout, while PPs report the largest proportion of patients with severe burnout.

The treatment practices depend on burnout severity. Psychiatrists and physicians with double specialty (GP-OPs and GP-PPs) treat patients with more severe burnout than GPs. Psychologists treating patients with severe burnout collaborate with other health professionals and contact the POB’s employer and/or health insurance. Treatment practices and burnout severity are not associated with the proportion of relapsed patients and patients who return to work. Yet, the former is associated with professionals’ age, sex, and specialty. Physicians with waiting time >3 months have a higher proportion of relapsed patients. GPs prescribe most often sick leaves, while PPs are the most frequent prescribers of pharmacological treatment. PPs collaborate significantly more often than GPs with pharmacologists and contact POB’s employer and health insurance. Among psychologists, work psychologists differ from other psychologists by a more frequent POB (psycho)education and coaching, namely on how to negotiate with employer and family, as well as on physical exercise. They also more often contact POB’s employer. Besides profession and specialization, we observed important regional variation in treatment modalities chosen by both physicians and psychologists.

## INTRODUCTION

Since May 2019, burnout is recognized as an occupational phenomenon resulting from “chronic stress at the workplace that has not been successfully managed” (1). In the 11th revision of the International Classification of Diseases (ICD-11), burnout is classified among “Factors influencing health status or contact with health services”, i.e., reasons for using health services, which are not ascribed as a disease (2). In its advanced stage, burnout shares several symptoms with depression, but burnout is mostly seen as a risk factor for depression or a mediator in the relationship between exposure to job stress and depression (3–5). Conversely to depression, established as a leading cause of disability worldwide and a major contributor to the overall global burden of disease (6, 7), burnout is recognized as a disease only in few countries (8). Despite a recently launched effort to harmonize the definition and assessment of occupational burnout (8, 9), no standardized and internationally accepted criteria are still available. Yet, as burnout strongly affects a large portion of population, especially in the health care sector (10–13), it was declared as a ‘public health crisis’ in the USA (14).

In Switzerland, the five sectors at highest risk of occupational burnout (i.e., sectors with the highest prevalence of job stress and emotional exhaustion) are: 1-banking and insurance, 2-health care, 3-security and safety, 4-transportation, warehousing and mail, and 5-accommodation and catering (15). In 2022, the Job Stress Index showed that the proportion of the Swiss professionally active who feel emotionally exhausted exceeded 30% (16). Indeed, since the COVID-19 pandemic, burnout is reported at unprecedented rates in all countries (17–19).

Considering this national and international context, our main objective was to explore the number, characteristics, and activities of Swiss health professionals who manage patients with occupational burnout (POB) in the Swiss outpatient sector.

In a prior qualitative study, we identified that health professionals who are the most concerned with POB in their clinical practice are general physicians (GPs), occupational physicians (OPs), psychiatrists-psychotherapists (PPs), psychologists, and occupational nurses (20). This study was set up to evaluate the resources available for POB management and to look at the current practices from a care quality and effectiveness perspective in order to identify uncovered needs and improvement potential. We thus aimed to address more specifically the activity of Swiss healthcare professionals in the outpatient sector with respect to POB and explore its potential determinants.

## METHODS

### Research questions and hypotheses

To reduce the risk of type I error, also known as a false positive, we limited our analysis to ten questions as follows: 1-Who among Swiss health professionals are confronted with POB in the outpatient setting? 2-Who treats the POB? 3-Who believes it is possible to cure occupational burnout, and who believes this is not possible? 4-Who has the highest proportion of patients who were able to return to work? 5-Who has the lowest proportion of patients who relapsed? 6-Does the stage of burnout severity influence the treatment choice? 7-Does the prognostic belief influence the treatment choice? 8-Does the proportion of relapsed patients depend on the stage of burnout? 9-Does the proportion of patients who relapsed depend on contacts with patient’s employer and health insurance physician or collaborations with other health professionals? 10-Does the proportion of patients who were able to return to work depend on contacts with patient’s employer and health insurance physician or collaborations with other health professionals? While the first five questions were purely descriptive, the other five were based on the pre-defined hypotheses to avoid data-driven results and enhance reproducibility. These hypotheses as well as the statistical analysis plan were determined prior to starting the analysis and are available at Unisante data depository, along with the protocol of this study (https://doi.org/10.16909/dataset/42).

### Study design and population

We applied the design of a cross-sectional study deployed throughout Switzerland using an electronic survey. The target population of the survey was Swiss health professionals who are concerned with the management of occupational burnout in the outpatient setting, i.e. physicians, psychologists and occupational nurses (Total n = 16’883). The lists and contact information for these professionals were obtained in collaboration with the Swiss Medical Association (FMH), the Swiss Federation of Psychologists (FSP), the Swiss Society for Psychiatry and Psychotherapy (SSPP), the Association of Organization and Work Psychologists (PSY4WORK.CH), and the Swiss Association of Occupational Health Nurses (ASIST). The study sample consisted of 3’216 health care professionals who participated between April 7 and July 20, 2021 in the electronic survey. Since the participation was completely anonymous, the need for consent was waived by the ethics committee (CER-VD BASEC-Nr. Req-2021-01156). Table 1 provides the number of respondents and response rates per profession and medical specialty.

**Table 1.** Reponse rate per profession and medical specialty in the STBOS-VD survey.

| **Profession** | **Target**<br>**population** | **Number of**<br>**respondents** | **Response**<br>**rate (%)** |
| --- | --- | --- | --- |
| Physiciens | 10 272 | 1 723 | 17 |
| General practitioners | 7 227 | 874 | 12 |
| Psychiatrists | 2 968 | 657 | 22 |
| Psychologists | 6 514 | 1 326 | 20 |
| Occupational health nurses | 97 | 39 | 40 |
| Other | - | 128 | - |
| Total | 16 883 | 3 216 | 19 |

**Table 2.** Descriptive characteristics of the study sample.

| Characteristics | N | % |
| --- | --- | --- |
| Total number of respondents | 3216 |  |
| Participants confronted to patients with burnout | 2951 |  |
| Participants treating patients with burnout | 1130 | 100.0 |
| Sex |  |  |
| Male | 414 | 36.6 |
| Female | 716 | 63.4 |
| Age |  |  |
| Less than 30 years | 17 | 1.5 |
| 30 - 39 years | 169 | 15.0 |
| 40 - 49 years | 306 | 27.1 |
| 50 - 59 years | 335 | 29.7 |
| 60 - 65 years | 157 | 13.9 |
| More than 65 years | 146 | 12.9 |
| Language of correspondance |  |  |
| French | 529 | 46.8 |
| German | 570 | 50.4 |
| Italian | 31 | 2.7 |
| Principal Swiss region |  |  |
| Région lémanique (VD, VS, GE) | 409 | 36.2 |
| Espace Mittelland (BE, FR, SO, NE, JU) | 228 | 20.2 |
| Nordwestschweiz (BS, BL, AG) | 138 | 12.2 |
| Zürich (ZH) | 157 | 13.9 |
| Ostschweiz (GL, SH, AR, AI, SG, GR, TG) | 97 | 8.6 |
| Zentralschweiz (LU, UR, SZ, OW, NW, ZG) | 70 | 6.2 |
| Ticino (TI) | 31 | 2.7 |
| Job category |  |  |
| Physician | 713 | 63.1 |
| Psychologist | 410 | 36.3 |
| Occupational Health Nurse | 2 | 0.2 |
| Other | 5 | 0.4 |
| Principal place of work |  |  |
| Private practice | 894 | 79.1 |
| Clinic or private care center | 60 | 5.3 |
| Hospital or public clinic | 121 | 10.7 |
| Public company | 13 | 1.2 |
| Private company | 22 | 2.0 |
| Insurance | 3 | 0.3 |
| Other | 17 | 1.5 |
|  | Median | IQR |
| Number of consultations in the last month | 85 | 50 - 150 |
| Number of consultations for burnout in the last month | 4 | 2 - 6 |
| Number of years in practice | 15 | 8 - 25 |

### Data collection

To collect the necessary data, a 48-item questionnaire was construed based on a first questionnaire developed by Droz and Wahlen (22) and the information obtained from our qualitative study described in previous reports (20, 21). The questionnaire was divided into four parts: 1-demographic data of the participants, 2-data on the burnout definition and detection, 3-data on the management and treatment of POBs, and 4-information on needs and suggestions for practice improvement. The full questionnaire as well as more details on its creation and testing were published previously (21).

A trilingual electronic version of the questionnaire was created using REDCap software and made available on the internet via a web address and a QR access code. Data entry fields had validation mechanisms and it was made sure that participants only participated once. The questionnaire was promoted through various means, including professional associations, press releases and a web page dedicated to the study. One reminder e-mail and a postal letter were sent to eligible professionals either by the professional associations or by the research team. The participation was voluntary and anonymous, the collected data were automatically transferred to a secure and access-restricted server.

Participants who were not confronted with burnout patients were asked to leave the questionnaire after their demographic data had been collected. Participants who did not manage burnout patients themselves were asked to leave the questionnaire before questions about treatment options were asked. The data did not contain any personal identifiers.

### Data processing and statistical analysis

First, we checked and recoded variables to reduce the number of categories. For instance, a new 7-class variable “Region of practice” was created: 1-“Lake Geneva region (VD, VS, GE)” 2-“Espace Mittelland (BE, FR, SO, NE, JU)” 3-“Northwestern Switzerland (BS, BL, AG)” 4-“Zurich (ZH)” 5-“Eastern Switzerland (GL, SH, AR, AI, SG, GR, TG)” 6. “Central Switzerland (LU, UR, SZ, OW, NW, ZG)” 7-“Ticino (TI)”. Furthermore, 18 dichotomic variables (yes/no) for options of burnout treatment were transformed in a 7-class variables “treatment options chosen”, for physicians and for psychologists, respectively. Finally, we dichotomized the outcome variables “Prognostic beliefs” as “Yes, burnout can absolutely be healed” versus “Burnout can sometimes or never completely be healed”; “Severity of burnout” as “Mostly mild or moderate burnout” versus “Mostly severe burnout”; “Proportion of patients with relapse” as “<= 25% patients who relapsed” versus “>25% patients who relapsed”; and “Proportion of patients fit for work” as “<= 75% patients that are fit for work” versus “>75% patients that are fit for work”.

The statistical analysis was conducted according to a pre-defined statistical analysis plan (SAP), where we defined each research question along with the statistical model and variables to answer it. This SAP was developed following a thorough descriptive analysis of the collected data and discussion with the project’s scientific committee (21). The SAP is available at the Unisante data depository (https://doi.org/10.16909/dataset/42)

We considered several datasets for model application. A full dataset with data from all respondents was used for the research questions 1 and 2. This dataset was then restricted to professionals who treat POBs and split by profession (i.e., physicians and psychologists). These datasets were used for the research questions 3 to 10, where the stratification by profession allowed us differentiating treatment options prescribed by physicians and therapies conducted by psychologists in the respective models.

As some questions in the survey allowed an answer option “Impossible to specify” or “I don’t know” and the same participants often chose these options multiple times (indicating the lack of a precise opinion), we excluded these options from the analysis. Keeping them would penalize the statistical performance and result interpretation by increasing the model complexity without adding any meaningful information. Similarly, the respondents with missing values on outcomes or covariates were excluded. The missing values represented less than 15% in average and mostly concerned the open questions rather than questions with multiple choice. The imputation technics in this case have limited or no efficacy.

For each research question we first carried out a univariable logistic or multinomial logistic regression model with the preselected dependent variable and predictor variable. In a second step, multivariable regression models were applied with all the independent variables of interest in the same model. The regression results were reported as Odds Ratio (OR) or Relative Risk Ratio (RRR) with associated 95% confidence interval (IC-95%) and p-value. Data-management procedures and statistical analyses were performed using STATA V 17.0 software.

## RESULTS

### Sample description

The sociodemographic and professional characteristics of responding professionals that treat occupational burnout are summarized in Table 1. The descriptive characteristics of physicians and psychologists are provided in Supplementary material Tables S1a and S1b, respectively. Among the 3216 respondents, 2951 reported having POBs, and 1130, including 713 physicians and 410 psychologists, treat them. Most (n=894, 79%) work in private practice, are German speaking and have 16.9-year professional experience on average. Among physicians (Table S1a), there was a similar percentage of men (48%) and women (52%) whereas among psychologists (Table S1b); there were 83% women. Among physicians who treat POBs, most were either GPs (47%) or PPs (48%), while among psychologists the majority were psychotherapists (82%).

### Proportion of consultations for patients with occupational burnout in professionals’ practice

The proportion of POB consultations was obtained by dividing the reported number of POB consultations by the total number of consultations (assessed over the last month of usual practice). This proportion enables approximation of the prevalence of POB among physicians’ psychiatrist’s clients. Indirectly, it might reflect the volume of outpatient clinical activities dedicated to POB among Swiss health professionals. We observed that this proportion varied according to the profession, medical specialty and psychological specialization (21). Among physicians, the reported proportion of POB consultations was 6.0%, but those with a double specialty (GP-PPs or OP-GPs) reported the highest proportion (10.8%), followed by OPs (8.7%) and PPs (7.3%). GPs’ reported proportion was 4.8% among their patients. Among psychologists (overall proportion of 8.6%), work psychologists reported the highest proportion (25.6%), while clinical psychologists and psychologists-psychotherapists reported significantly lower proportion of POB among their clients (8.9% and 6.9%, respectively). The reported proportion of POB consultations also differed by professionals’ region of practice (Figure 1).

**Figure 1.**
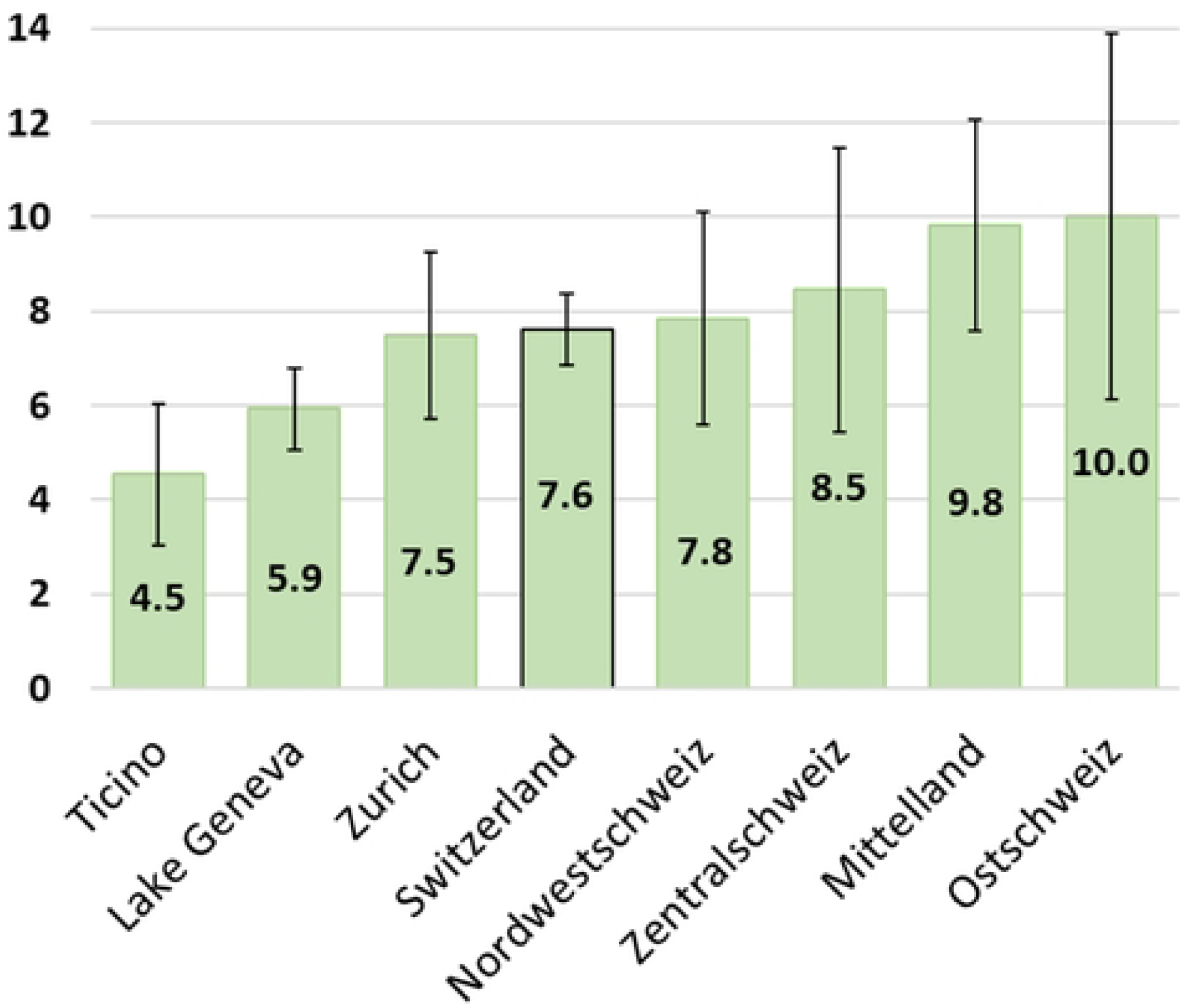
Average proportion of consultations for patients with occupational burnout (mean and 95% confidence interval) in the Swiss health professionals’ activity during the last month (STOBS-VD survey, June 2021)

### Stage of burnout severity and prognostic beliefs

Among the major groups of professionals, PPs had the highest proportion of patients with severe burnout (49%) (Figure 2). Psychologists had lower proportions of patients that relapse than physicians but, in many cases, could not specify it. In contrast, the proportions of patients that could return to work were similar for physicians and psychologists when omitting those unable to specify this. As for the prognostic beliefs, the proportion of physicians and psychologists who believe that occupational burnout can absolutely be cured were similar (around 51%); yet, GPs had a slightly more optimistic prognostic belief.

**Figure 2.**
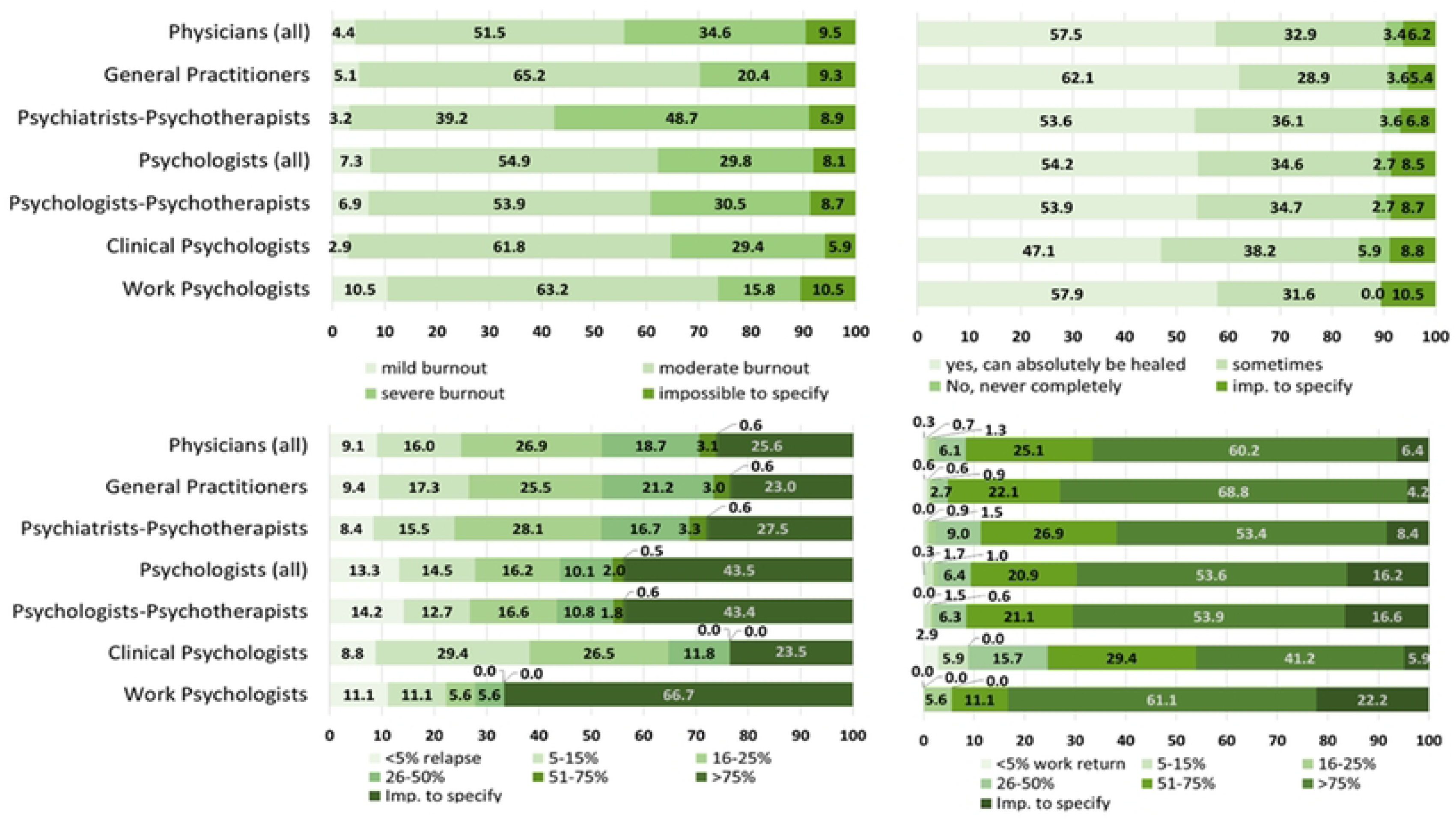
Distribution (in %) of health professionals per studied outcomes: A – Stage of burnout severity in majority of patients/clients; B – Believes on the prognosis of burnout; C – Proportion of treated patients/clients who relapse; D-Proportion of treated patients/clients who can return to work.

These distributions differed by geographic region, as demonstrated in Supplementary Material Figure S1. While professionals in the Mittelland region reported the lowest proportion of severe POBs, they reported more POBs consulting them than physicians in other regions, except the Ostschweiz (Figure 1). On the other hand, professionals in Ticino report the highest and those in the Lake Geneva Region the second highest proportion of mainly severe BOP.

### Professionals’ profiles with respect to burnout treatment and treatment outcomes

The results relative to the research questions 1 to 5 are provided in Supplementary materials Tables S2 to S9, either for all participants or stratified by profession. Regarding the question 1, we found that psychologists had a lower probability to be confronted with POBs at the first instance than physicians [OR=0.55, p=0.003] (Table S2a). However, psychologists seem to treat POBs more often than physicians [OR=1.97, p<0.001] (Table S3a), especially psychologists specialized as psychotherapists [OR=6.35, p<0.001] (Table S3c). Among physicians, psychiatrists treat burnout more frequently than GPs [OR=12.6, p<0.001] (Table S3b). Since neither age, sex, the number of consultations in the previous month nor the number of years of practice were associated with the treatment of occupational burnout (Tables S3a-c), no profile emerged as response to the research question 2.

Regarding the research question 3, we found that some more pessimistic beliefs were associated with 60+ age in physicians [OR 2.2, p=0.011] (Table S4a) and female sex in psychologists [OR 2.15, p=0.024] (Table S4b). German-speaking professionals (both psychologists and physicians) appeared less optimistic regarding burnout prognosis than French speaking professionals, although the regional variation was more pronounced among psychologists than among physicians.

As response to the research question 4, we found that compared to GPs, other medical specialists treating POBs reported lower proportion of patients able to return to work. This was also the case of physicians whose waiting time for consultation appointment is longer than one month (compared to physicians with a sooner consultation possibility) and of physicians practicing in the regions of Zurich, Ostschweiz and Ticino (compared to physicians in Leman region) (Table S5a). In contrast, physicians practicing in the Zentralschweiz region reported a higher proportion of patients able to return to work than Leman physicians. Among psychologists, we observed no regional variation (Table S5b). Compared to clinical psychologists, psychologists-psychotherapists reported significantly more often a high proportion of clients who returned to work. Psychologists aged between 40 and 59 years old, had also a higher proportion of such clients that younger psychologists.

Regarding the research question 5, we found that in both physicians and psychologists, 60+ age and female sex were associated with a lower probability of relapse among the treated POBs. In contrast, a longer waiting time for consultation appointment, was associated with a higher chance to have more relapsed patients after treatment among physicians (Table S6a). Among psychologists, an opposite association was observed (Table S6b).

### Factors associated with the choice of treatment modalities

As expected, the choice of treatment modalities was highly determined by professional specialty or specialization (Figure S2). GPs prescribe most often sick leaves, while PPs are the most frequent prescribers of pharmacological treatment. PPs collaborate significantly more often than GPs with pharmacologists and contact POB’s employer and health insurance. Among psychologists, work psychologists differ from other psychologists by a more frequent POB (psycho)education and coaching, namely on how to negotiate with employer and family, as well as on physical exercise. They also more often contact POB’s employer. Besides profession and specialization, we observed important regional variation in treatment modalities chosen by both physicians and psychologists (Tables S7a and S7b). Physicians in Ticino and in the German-speaking regions were more prone to contact POB’s employer or health insurance doctor in addition to the treatment prescription than in the Leman region. This was especially the case of OPs, PPs, and physicians with double specialty.

The probability for physicians to contact POB’s employer as part of the treatment procedure was associated with a more pessimistic belief on burnout prognosis (RRR=1.78, 95%-CI=1.20-2.62), but after adjustment for other covariates, the association turned to a borderline statistical significance (RRR=1.48, 95%-CI=0.97-2.26) (Table S7a). Similar associations were observed for a combination of the treatment prescription, employer contact and interdisciplinary collaborations, however, it was not associated with practitioner’s belief on burnout prognosis. In psychologists, a pessimistic belief on burnout prognosis was also associated with the combination of therapy and contact with the POB’ employer, with a similar association strength (RRR=1.81, and 1.63, before and after adjustment, respectively) than among physicians (Table S7b). The combination of therapy with employer and/or health insurance contact and collaboration with other professionals among psychologists was twice more frequent for clients with severe burnout (Table S8). In physicians, the stage of burnout severity was associated with employer contacts and interdisciplinary collaborations only in univariable analysis.

In response to the research question 8, we found that the proportion of relapsed patients reported by respondents does not depend on the stage of burnout severity. The results were in opposite directions in physicians and psychologists, with OR=0.84 (95%-CI=0.56-1.27) and OR=1.69 (95%-CI=0.88-3.26), respectively. The results corresponding to the research question 9 and 10 are provided in the Table S9. We found no association between interdisciplinary contacts or collaborations and the proportion of relapsed patients reported by healthcare professionals.

However, the association between the collaboration with pharmacologist and proportion of patients who were able to return to work was negative and statistically significant, meaning that professionals seek pharmacologist help when their patients cannot manage to return to work.

## DISCUSSION

### Main findings

The study provided a state of the art on the number and distribution of Swiss professionals confronted with POBs, the volume of consultations dedicated to management of POBs in their practice, the severity of burnout in their POBs, their general beliefs on burnout prognosis, and treatment options chosen including interdisciplinary collaborations and collaboration with POB’s employer and health insurance. The latter was particularly interesting since the Practical recommendations on burnout treatment released by the Swiss network of experts on burnout highlight the importance of interdisciplinarity in the POB treatment (4, 5). The study showed that POB management constitutes 5 to 25% of health care professionals’ outpatient consultations, with an inequal distribution of POBs across professionals’ specialties and specializations, but also across geographic regions. The latter might raise an issue of health inequality if the Swiss POBs cannot receive the same health care depending on where they live. This raises in turn a crucial issue of perception and measurement of occupational burnout across regions and cultures in Switzerland.

Indeed, regional variation was the most salient and consistent finding in most research questions, especially those aimed to characterize professionals’ profile (Questions 1-5). The regional variation in practice can originate from several sources including regional differences in professionals’ initial and continuous education; disparities in health organization and healthcare services availability and accessibility; cultural differences in perception of mental illness and occupational burnout as well as their (de)stigmatization; and influence from border countries. The latter was pointed out in health insurance statistics, according to which sick leaves are significantly more frequent and longer in the French-speaking Switzerland (20, 21). Such regional variations in professionals’ practice are also remarkable since the Practical recommendations on burnout treatment exist in Switzerland since 2016 (4, 5), and probably reflect professionals’ individual sensitivity about the problem and its management.

We observed that the stage of burnout severity determines the treatment choice as does the health care professionals’ beliefs on the burnout prognosis. Physicians and psychologists tend to add more collaborative actions and contacts with POB’s employer and/or health insurance mostly for severe POBs and a pessimistic prognosis. This echoes the professionals’ identified gap in the current practice and the need of tripartite collaboration (POB, employer, health professionals) as early as possible (23–25).

All these results seem logical, however by objectivating them using a large and representative study sample, a comprehensive questionnaire developed by a multidisciplinary research team, and a rigorous analytical scheme, these results are the first to translate empirical guesses on burnout management in Swiss outpatient sector into scientific knowledge (26). For this, some abstractions were still necessary, namely the outcome variables allowing to compare and to look at the current practices from a care quality and effectiveness perspective. These abstractions deserve a careful definition and interpretation, with consideration of the study context.

### Relevance and accuracy of the studied variables

We defined four outcomes to analyze the professionals’ profiles and practices. The stage of occupational burnout was defined using a set of open questions on how the professional defines the burnout and each of its stages of development/severity, considering three main stages: mild or early burnout, moderate burnout and advanced or clinically severe burnout. The responses were translated and analyzed using the thematic context method in MaxQDA. The resulting definitions were consistent across professions and regions (21), thus we are confident that respondents correctly evaluated this outcome for most of their POBs.

Regarding the belief on the burnout prognosis, an average assessment for most of one’s POBs can be more challenging, as prognosis depends on multiples factors such as the etiology, number and severity of symptoms, therapeutical alliance, patient’s treatment compliance and collaboration, as well as availability of the others’ support and collaboration. We observed, however, that most respondents could assess and report their general prognostic beliefs, and that the rate of those who could not specify it was even lower that for the stage of burnout (Figure 2).

Conversely, the proportion of relapsed patients among the treated POB raised a concern, since for a quarter of physicians and more than 40% of psychologists, especially the work psychologists, it was impossible to precise the relapse rate (Figure 2). The discussion with the scientific committee helped contextualize it, to understand the reason and the impact on the study results. Indeed, one reason is that a burnout relapse is rarely announced to the professional by the patient. Moreover, in a psychotherapy context, a patient who relapsed may tend to change his/her psychotherapist, either following a misalliance with the therapist or for seeking a new or different treatment approach. Therefore, the accuracy of this outcome may be an issue. Since we excluded those who could not estimate the average rate of relapse in their patents/clients, the remaining subsample is likely to be too small to yield results with sufficient statistical power. This can explain why most associations with this variable are statistically non-significant and challenging to interpret.

Finally, the average rate of POBs ability to return to work after the treatment was well assessed. Return to work generally symbolizes the recovery (27) or a “return to the normality” (28). Since return to work is likely to be an outcome, seen as the therapy effect, it can also correspond to the end of the treatment when return to work seems successful (29, 30). Therefore, it is relevant to assess and to examine this outcome in relation with the treatment modalities as we did.

Regarding the treatment options, in this study we focused on a combination of usually prescribed treatments or therapies with the contact of POBs’ employers, health insurance and collaborations with other health professionals. For this, we abstracted a 7-class aggregated variable from 18 options that respondents could select in a multiple-choice question. By doing this we collapsed the details on therapies prescribed and conducted (Figure S2), as their consideration without individual POB data would be meaningless. In contrast, it would be relevant and interesting to consider the different combinations of treatment options delivered at an individual POB and professional level. This was not possible in the frame of this study but could be feasible in a prospective cohort study of POBs.

### Contextual relevance of study findings

It is noteworthy that this study was initiated just before the Covid-19 pandemic. The survey questionnaire was distributed when the preparation of the 1^st^ vaccination campaign started, which explains that some cantonal associations of GPs and family doctors refused to collaborate and promote this study. This also can explain the relatively low participation rate, although it is comparable to other surveys conducted in the field before or after the pandemic (34). However, as the Covid-19 pandemic increased the incidence and prevalence of mental ill-health generally and of burnout particularly (11, 19), the proportion of consultations for POB estimated in this study is likely to under-represent the post-pandemic reality. The pandemic might also have increased the proportion of severe POBs, the relapse rate, and have decreased the proportion of those who are able to return to work, as the work conditions worsened in many instances (6, 35). Consequently, the study findings reflecting the pre-pandemic situation cannot be directly extrapolated to the pandemic and post-pandemic situation and should be confirmed in an evolved context.

The new national regulation authorizing psychologists-psychotherapists to practice within the framework of compulsory health insurance independently from psychiatrists is another contextual change to mention. This regulation entered in operation in July 2022 and might change the figures on outcome distribution across professions and regions observed in the present study. This could be another reason for repeating this study in a near future and check the result reliability in a new context.

### Study limitations

This study has several limitations that deserve discussion. Its cross-sectional design is suitable for producing a reliable picture of the health care professionals in outpatient sector dealing with POB and describe their activities in general but precludes a formal comparison of their practices with the Practical recommendations on burnout treatment released by the Swiss network of experts on burnout (4, 5). Therefore, such a comparison was beyond the scope of the present study but could be valuable in the future. The 19%-overall response-rate was rather modest but that of GPs (12%) was particularly low. Given the Covid-19 pandemic context, such a low participation may conduct to a self-selection of healthcare professionals the most concerned with BOP management. In turn, this may lead to an over-estimation of POB proportion and a biased description of practice distributions. As we could only assess the outcomes on an aggregated level (i.e., as an average for most professionals’ POBs), some of observed associations can be ecological and need confirmation in future studies based on individuals-level data. As far as we know, such studies of POB do not exist in Switzerland and are also extremely seldom abroad (31–33). Such studies will also enable comparing the effectiveness of different treatment options and their combination and producing or updating practice recommendations regarding burnout management.

Although we limited the number of statistical analyses, multiple comparisons using the same dataset may be still an issue, leading to an increased likelihood of finding significant results by chance. Yet, the result interpretation and validation were conducted in collaboration with the project scientific committee members to stay true to the original research questions and hypotheses and enhance the validity and credibility of the study.

## CONCLUSION

This study is the first to objectify the importance of occupational burnout as a health care need in the outpatient sector in Switzerland. In particular, the study findings point out its relative importance in general practitioner practice and a potential interest of their training as initiators and coordinators of burnout treatment. The study revealed that most professionals collaborate with other health professionals and patient’s employer and health insurance only in cases of severe burnout, pessimistic beliefs on burnout prognosis, and when the return to work is challenging. The support of this collaborative, interdisciplinary efforts and their extension to earlier burnout stage might thus improve the treatment outcomes. Education regarding occupational burnout detection and management seems also necessary, especially in GPs pre-graduate curriculum. As the study was cross-sectional with some aggregated outcomes, its findings should be considered with caution and confirmed in future studies, ideally using prospective cohort design and individual burnout patient data.

## ETHICAL CONSIDERATIONS

The study protocol was submitted to the cantonal ethics commission for Research on Human Beings of the Canton of Vaud (CER-VD). The CER-VD judged that this study did not fall within the scope of the Federal Law on Research on Human Beings (HRA) and issued an exemption from review (BASEC-Nr. Req-2021-01156).

## AUTHOR CONTRIBUTIONS

IGC, ND, and ND conceived the project and obtained funding. IGC coordinated the study and drafted the manuscript. RG performed data management and statistical analysis and prepared figures and tables of study results. All co-authors contributed to the questionnaire development or testing, survey dissemination, result interpretation and critical review of the manuscript. All authors agreed on the final version of the manuscript.

## Data Availability

All relevant data are within the manuscript and its Supporting Information files

## ACKNOWLEDGEMENTS

We thank persons and organization who help prepare and distribute the survey and supported this study: S. Blanc, O. Talpain, J. Banrejee, A. Farine, E. Plys, M. Al Gobari, T. Charreau, and B. Chiarini from Unisanté; S. Barras Duc (FOPH); A.S. Brandt (SECO); M. Brinkrolf (FSP); M. Dominicé Dao and I. Guessous (HUGe); M.T. Giorgio (SSMT); B. Weil (FMH); Medical associations of the cantons of Aargau, Bern, Geneva, Glarus, Jura, Neuchatel, Solothurn, St. Gallen, Thurgau, Vaud, and Wallis. We also thank all respondents for their time and participation in the study.

## SUPPLEMENTARY MATERIAL / DATASETS

1 file with 18 tables and 2 figures

